# Appropriate intensification of diabetes-related therapy by general practitioners: cross sectional study

**DOI:** 10.1101/2020.04.07.20057380

**Authors:** Nelson Aguirre, Peter Carswell, Tim Kenealy

## Abstract

**Aim:** To measure intensification of therapy (the inverse of clinical inertia) by general practitioners in the management of hyperglycaemia, hypertension and dyslipidaemia in patients with diabetes.

**Methods:** We analysed clinical practices results for Hb1c, blood pressure, lipids and pharmacy records for prescriptions according to the guidelines. We identified whether medication was increased at each patient visit when the goals for glycosylated haemoglobin (<= 64 mmol/mol), systolic blood pressure (<=130 mmHg) and LDL cholesterol (<=3.5 mmol/L) had not been reached at the time of the visit. The study was conducted between January 2009 and December 2011 in South Auckland, New Zealand.

**Results:** Data from 314 patients with diabetes included 2398 prescriptions. Appropriate intensification of therapy was seen in 31% of 457 visits when HbA1c was raised, 11% of 375 visits when systolic blood pressure was raised and 2% of 51 visits when LDL cholesterol was raised.

**Conclusions:** There are clear differences in intensification therapy in terms of the co-morbidity addressed. Clinical inertia may be one important and changeable contributor to diabetes complications. New Zealand has replaced “get Checked” programme for Diabetes Care Improvement Package, which is a primary care based programme seeking to improve outcomes for people with diabetes. This new programme is based on coordination of diabetes care, therefore a further analysis measuring clinical inertia may show improvements in quality, consistency and direction of care for people with diabetes in New Zealand.

**Highlights:**

- We measure intensification of therapy by general practitioners.
- Management of hyperglycaemia, hypertension and dyslipidaemia were tested.
- There are differences in intensification therapy in terms of each co-morbidity.
- Clinical inertia may be one changeable contributor to diabetes complications.

## INTRODUCTION

Chronic diseases such as diabetes can have devastating consequences on morbidity, mortality and cost (1, 2). The prevalence of diabetes and related diseases is increasing worldwide(3-5). There is a vast amount of clinical evidence suggesting the benefits of the glycaemic control (6, 7), however more than 40% of these patients struggle to reach those targets (8, 9).

Management of type 2 diabetes in New Zealand is largely undertaken in primary care, with diabetes specialists involved in care of more complex patients and in support of primary care providers.(10) General practitioners (GPs) act as a central contact point for patients with type 2 diabetes and share, with patients, the primary responsibility for achieving optimal outcomes.(11-13) Nevertheless GPs will sometimes fail to increase therapy when it appears to be indicated, action that is recognised as clinical inertia.(14-17)

Observational studies in the U.S, Canada and Europe have shown rates of clinical inertia between 25 % and 83 %.(16, 18) Several different methods have been proposed to measure clinical inertia.(16, 17, 19) In the case of diabetes, measurement should go beyond pharmaceutical management of glucose control to include therapy for blood pressure and cholesterol.(20) Clinical inertia may one of the reasons why patients with diabetes do not reach management goals. Measuring intensification of therapy as a the first steps to understand the size of the problem and perhaps point to ways to support GPs managing patients with diabetes.(17)

The primary goal of this study was to measure the percentage of clinical inertia over the time in response to the clinical needs of the patients, in a region with high prevalence of diabetes in New Zealand.

## METHODS

### Context

Counties Manukau in South Auckland, New Zealand, had about 470,000 residents in 2011, of whom 8% of the adult population, accounting for 14% of people with diabetes in New Zealand.(21, 22) Health care is funded by a tax-based system in which secondary care is free to the patient, and primary care and pharmaceuticals are heavily subsidised. “Get checked” programme was a diabetes follow up care fully funded in New Zealand which ceased on July 2012 after one decade. The programme gave diabetes patients access to a free annual health check, including key tests and prescriptions based on planning for the year ahead.(21).

### Data Sources

Data were provided by a Primary Health Organisation (PHO). Measures of blood pressure, HbA_1c_ and LDL cholesterol (clinical data) came from data submitted by general practices to the PHO for payment under the annual review programme. Prescription data was collected directly by the PHO from community pharmacies; all the prescriptions were already dispensed by patients. The PHO was not able to collect data in all the practices, as they used different software and formats. In other cases, general practice managers did not allow downloading of their prescription databases.

### Study Subjects

We invited all practices affiliated with the PHO, located in Counties Manukau, who had submitted diabetes annual reviews for patients with type 2 diabetes. This included more than 140 GPs in 80 general practices. Patient inclusion criteria were: type 2 diabetes; at least two primary care visits during the 6 months period beginning Jan 2009; a measurement for at least one of HbA_1c_, systolic blood pressure or LDL cholesterol in the 18 months prior to their first included visit; and at least one primary care visit between 1 and 11/2 years after the index visit.

### Analysis

We considered each medication dispensing as evidence of a medical decision that this medication was appropriate for this patient at this time. We assessed each dispensed medication against data assumed to be available to the prescribing doctor – blood pressure measured on the day of dispensing or earlier, and HbA1c & cholesterol tested / result date (which) one day or more prior to dispensing, or earlier. In cases where no clinical data were available since the previous prescription, no judgement about appropriateness could be made.

Schmittdiel et al. defined intensification of treatment as meeting one or more of three criteria: an increase in the number of drug classes; an increase in the daily dosage of at least one ongoing drug class; or a switch to a medication in a different drug class. We grouped medications into classes: seven classes for blood pressure (ACE inhibitors, angiotensin antagonists, beta adrenergic blockers, calcium channel blockers, thiazides/related diuretics, potassium-sparing diuretics, and loop diuretics); five class for hyperlipidaemia (statins, bile acid resins, fibrates, niacin, and ezetimibe), and four class for glucose control (sulfonylureas, metformin, thiazolidinedione, and insulin); combination pills were considered as consisting of two classes.(23) Novel agents such as GLP-1s, and DDP VI were not included in the study as they are not part of the diabetes management in primary care in new Zealand.

Target values for HbA1c (<64), systolic BP (<130) and LDL (<3.4) were determined from target values used in previous studies of clinical inertia(16, 17, 20, 24, 25) and from New Zealand cardiovascular and diabetes management guidelines at the time.(26) Participating GPs considered these targets as appropriate (semi-structured interviews not reported here). A value above target that did not result in a change of medication at the next prescription was taken as evidence of clinical inertia, or failure to intensify therapy.

Intensification of treatment was calculated for each clinical category (glucose, blood pressure and cholesterol) as the number of prescriptions that showed intensification divided by the number of prescriptions.

Data management and analysis of the data were performed using Stata 11.1 for Windows (Stata, College Station, Texas).

### Ethics

All participating GPs provided written informed consent. Patients with diabetes were not identifiable all data were anonymised by the PHO. Ethics approval was obtained from Northern X Regional Ethics Committee reference NTX/11/EXP/150.

## RESULTS

The study sample consisted of 314 patients with diabetes, managed by 14 GPs over 2,398 prescriptions. All patients had diabetes, and most also had antihypertensive therapy (76%) and antilipemic therapy (62%). Table 1 summarises patients’ clinical data and numbers and clinical management.

**Table 1.** Characteristics of 314 patients with type 2 diabetes

| Characteristics | Number / (%) |
| --- | --- |
| Clinical Data |  |
| HbA1c mean (mmol/mol) | 68.2 ± 21.5 |
| Systolic Blood Pressure mean (mmHg) | 134 ± 17.9 |
| LDL mean (mmol/L) | 2.27 ± 0.77 |
| Prescriptions by type of drug | 2,398 |
| Antidiabetic (oral and insulin) | 1033 (43%) |
| Antihypertensive | 812 (34%) |
| Antilipidemic | 553 (23%) |

### Management of goals

All patients with diabetes were prescribed with oral drugs and/or insulin during the study period. A total of 78 from 314 (25%) patients and 268 from 2,398 (11%) prescriptions involve insulin. A change in medication therapy was considered necessary at 37% of all visits overall, shown by therapeutic category in Table 2.

**Table 2.** Intensification of therapy

| Characteristics | Number / (%) |
| --- | --- |
| Visits where HbA1c above 64 mmol/mol | 467 (45%) |
| Prescriptions showed Intensification of therapy | 144 (32%) |
| Visits where systolic blood pressure above 130 mmHg. | 375 (46%) |
| Prescriptions showed intensification of therapy | 42 (11%) |
| Visits where LDL cholesterol above 3.5 mmol/L | 51 (9%) |
| Prescriptions showed intensification of therapy | 1 (2%) |

An average intensification percentage, per condition, was calculated for each GP. Table 3 shows how is the distribution of the intensification therapy per general practitioner in each level of measure.

**Table 3.** Intensification Therapy by GP (n=14)

| GP code | % Intensification of Therapy |  |  |
| --- | --- | --- | --- |
|  | Glucose<br>% (denominator) | Blood pressure<br>%(denominator) | Cholesterol<br>% (denominator) |
| GD00114 | 33% (12) | 11% (9) | 100% (1) |
| GD00120 | 50% (4) | 0% (3) | 0% (3) |
| GD00310 | 54% (26) | 9% (46) | 0% (2) |
| GD00312 | 31% (13) | 10% (10) | 0% (3) |
| GD00330 | 42% (26) | 0% (9) | 0% (3) |
| GD00335 | 38% (56) | 7% (57) | . |
| GD00427 | 12% (90) | 10% (10) | 0% (11) |
| GD00512 | 36% (137) | 20% (94) | 0% (16) |
| GD00514 | 24% (33) | 13% (23) | 0% (2) |
| GD00535 | 43% (21) | 4% (49) | 0% (4) |
| GD00630 | 0% (5) | 0% (5) | . |
| GD00731 | 11% (18) | 10% (20) | . |
| GD00926 | 33% (15) | 8% (12) | 0% (1) |
| GD00930 | 36% (11) | 14% (28) | 0% (5) |

## DISCUSSION

Rates of clinical inertia shown by GP prescriptions for patients with type 2 diabetes in Counties Manukau range were 69% for glucose control, 92 for blood pressure and 91% for cholesterol. There was considerable variation between GPs. These rates of clinical inertia could substantially contribute to diabetes complication rates. We note that clinical inertia measured in this way, while consistent with the literature, does not allow for the fact that changes of therapy are negotiated with patients, and there will be times when practitioner and patient agree to assess the effect of lifestyle modification before increasing pharmaceutical management.

This study is consistent with findings in previous research in primary care.(18, 27, 28) Van Bruggen et al reported clinical inertia in diabetic patients as 43% for glucose control, 63% for blood pressure and 90% for cholesterol. Also in diabetic patients, Rodondy et al reported rates 34%, 36% and 44% respectively. In each case the clinical inertia rates are in the same rank order as for our study. This is likely to reflect the ongoing disproportionate emphasis on glucose control that is typical internationally.

This study describes proportions of clinical inertia rather than exploring causality. Explanations in the literature included physician’s overestimation of their adherence to guidelines, acceptance of elevated risk factors levels in their patients, and low motivation to treat asymptomatic chronic conditions.(29-31) Systematic attempts to improve rates of clinical inertia have been relatively intensive and may not be easily reproduced in routine practice. One example is how reminders and feedback from endocrinologist in secondary care to GPs about intensification of therapy could improve prescription behaviour. (32) However a more holistic approach taking in account health system barriers, the provider, and patient is needed in order to have a better understanding of this issue and implement better strategies to increase the intensification therapy. (33)

The Ministry of health made the decision to stop the “get Checked” programme based on the results after 10 years. One of the reasons was the marginal clinical improvements for people with diabetes in new Zealand. (34) The current study supports those findings, in the way that clinical inertia is a relevant issue that may explain the disappointing clinical outcomes on this population.

There are several limitations to our study. The GPs who agreed to contribute data may not be representative of other GPs. Given that study volunteers are generally considered to be among the better performers, this may lead to an underestimate of clinical inertia. We noted earlier that intensification of therapy includes agreement with the patient, not accounted for in our measurements, which may lead to us over-estimating clinical inertia. A small proportion of prescriptions for diabetic patients come from secondary care or after-hours clinics rather than from the patient’s usual doctor. Where blood pressure and laboratory results were not available between prescriptions it was not possible to adjudicate on clinical inertia, and it is unclear which direction this might affect our estimates. We used dispensing data (what the patient collects at the pharmacy) rather than prescribing rates (what the GP prescribes). There are times when a patient is given a prescription but does not collect the medication, perhaps due to cost. In a similar population we have previously shown about a 7% difference between prescription and dispensing.(35)

Despite these shortcomings, we have shown rates of clinical inertia to be sufficiently high to impact on diabetes complication rates. It therefore warrants further study of causation and evaluations of efforts to address it. New Zealand has replaced “get Checked” programme for Diabetes Care Improvement Package, which is a primary care based programme seeking to improve outcomes for people with diabetes. This new programme is based on coordination of diabetes care, therefore a further analysis measuring clinical inertia may show improvements in quality, consistency and direction of care for people with diabetes in New Zealand.

## Data Availability

No data is available

## Acknowledgements

We would like to thank all General Practitioners who took part in the study. And Paul Roseman and his team in ProCare PHO for support and assistance during the data collection.

